# Renal function is preserved following Tenofovir Disoproxil Fumarate (TDF) initiation among Rwandan’s living with HIV

**DOI:** 10.1101/2020.05.27.20114249

**Authors:** Gallican N. Rwibasira, Hae-Young Kim, Christella Twizere, Donald R Hoover, Qiuhu Shi, Adebola Adedimeji, Jonathan Ross, Gad Murenzi, Jean d’Amour Sinayobye, Kathryn Anastos

**Affiliations:** Rwanda Military Hospital, Kigali, Rwanda; Department of Public Health, New York Medical College, Valhalla, NY, USA; Centre National de Référence en matière de VIH/SIDA, Bujumbura, Burundi; Department of Statistics and Institute for Health, Health Care Policy and Aging Research, Rutgers The State University of New Jersey, New Brunswick, NJ, USA; Albert Einstein College of Medicine, Bronx NY, USA

**Keywords:** Tenofovir, renal function, Treat All, HIV

## Abstract

**Background:** Tenofovir disoproxil fumarate (TDF) is the antiretroviral drug most commonly associated with renal dysfunction. However, few studies have examined this association in sub-Saharan Africa despite recent scale-up of antiretroviral therapy (ART) to all people living with HIV (Treat All) in this region. We assessed estimated glomerular filtration rate (eGFR) change among HIV infected Rwandan adults following first line TDF-based therapy initiation.

**Methods:** This prospective, observational study was conducted in 10 Rwandan health centers. Participants were ART-naive adults (≥18 years) living with HIV who initiated TDF-based ART from 1^st^ July 2016 through 30^th^ July 2018. The primary outcome was eGFR change from pre-(within 12 months) to post-TDF initiation (within 6 months).

**Results:** Of 476 patients with pre- and post-TDF eGFR measurements, 264 (55.5%) were women and mean age was 35.9 years (SD 9.6). Mean pre-TDF eGFR was 92.4 (SD 24.0) and mean post-TDF was 96.0 (SD 21.0) mL/min/1.73m^2^. Mean pre-to post-TDF change thus increased 3.60 (SD, 26.6) mL/min/1.73m^2^ (p=0.001).

**Conclusion:** We detected a statistically significant clinically small renal function improvement within 6 months following TDF initiation among 476 ART-naïve patients. This supports continued TDF use for first-line treatment.

## Introduction

Tenofovir disoproxil fumarate (TDF) is a well-tolerated, once-daily antiretroviral (ARV) first-line HIV-infection therapy. Multiple studies conducted primarily in Europe and North America, observed long-term TDF use to be associated with proximal tubular dysfunction and impaired renal function (1),(2)(3) with increased risk of renal impairment especially among patients with advanced age, chronic kidney disease, longer TDF treatment duration, or treatment with ritonavir-boosted protease inhibitors.(1)(4)(5),(6)

Relatively few sub-Saharan Africa (SSA) studies have examined the association between TDF and renal dysfunction and revealed that TDF is associated with significant increase in renal impairment after long period of therapy (2). However, other African studies revealed conflicting results. Of the few studies from SSA, most have reported low incidence of TDF-associated renal dysfunction among PLWH initiating combination antiretroviral therapy (cART). However, in some studies TDF was associated with declines in eGFR (7) as well as Fanconi syndrome^1^. Furthermore, the efficacy of TDF as part of cART has been demonstrated in various studies(8)(9). Though TDF was recently proven to be effective and associated with few side effects (7)(8), further studies were recommended to support the continuous use of TDF in first line therapy(9)

Given TDF’s efficacy in reducing HIV/AIDS-related mortality(10)(1), its use is increasing widely with the adoption of Treat All guidelines by nearly every country in SSA(2)(3) To our knowledge, however, no studies have reported on the association between TDF and renal dysfunction under Treat All despite the scale up of TDF as part of first-line ART in SSA and the conflicting results reported from this region on TDF-associated renal dysfunction. We thus aimed to evaluate prevalence and predictors of TDF-associated renal impairment among people living with HIV (PLWH) following Treat All implementation in Rwanda.

## Materials and Methods

### Study design

A prospective, observational study using data from ten Rwandan health centers that participate in the Central Africa International Epidemiologic Databases to Evaluate AIDS (www.iedea.org) was conducted. CA-IeDEA is a multi-country project collecting secondary data from patients receiving HIV care and treatment in the Central African region; one of seven regions that comprise a global IeDEA network (www.iedea.org).This study was approved by the Rwanda National Ethics Committee and the institutional review board of the Albert Einstein College of Medicine.

### Setting and population

In July 2016, Rwanda, an East African country with 12 million persons and an overall 3% HIV prevalence(11), became one of the first countries to implement Treat All nationally. Under this policy, all persons living with HIV (PLWH) are referred for ART immediately upon diagnosis after performing a routine serum creatinine test to determine regimen choice (12). Current ART guidelines include TDF as a component of the main first-line treatment and recommend a serum creatinine measurement prior to initiating ART, after one month of use, and then yearly while on TDF. For this analysis, we included adults (≥18 years receiving HIV care at CA-IeDEA affiliated health centers who initiated a first-line TDF-based regimen from 1 July 2016 through 30 July 2018, and whose serum creatinine was measured both before (within 12 months) and after (within 6 months) of TDF initiation.

Patients with diabetes and/or hypertension, those on second-or third-line ART, and those who did not have creatinine measured both before and after TDF initiation were excluded.

Among 1,536 patients who initiated TDF during the study period, i) 205 (13.3%) did not have any creatinine measurements within 12 months before or 6 months after starting TDF, ii) 456 (29.6%) had only pre-TDF creatinine measurements, and iii) 399 (22.0 %) had only post-TDF creatinine measurements. Thus iv) 476 (31.0 %) patients had both pre- and post-TDF creatinine measurements. A comparison of these above four groups demonstrated statistically significant differences in proportions who were women, and mean age, CD4 count, creatinine and eGFR. Details are provided in the **Supplemental Table** and **Supplemental Figure**.

### Outcomes and predictor variables

The primary outcome was change in eGFR after initiating TDF. We considered pre-TDF creatinine as the closet measure within one year prior to initiating TDF, and post-TDF as the first post-TDF initiation measurement within one to six months after initiation.

Serum creatinine was measured in mg/dl. The CKD-EPI equation estimated GFR: eGFR = 141 x min (SCr/κ, 1)^α^ x max(SCr /κ, 1)^-1209^ x 0.993^Age^ x 1.018 [if female] where κ = 0.7 if female and 0.9 if male and α = −0.329 if female and −0.411 if male. The North American adjustment for sub-Saharan African race was not used as this adjustment has been shown not to apply within Africa itself and moved the estimates in our population towards the non-normal range. We also examined the change in creatinine after initiating TDF. Additional baseline variables evaluated here included sex, age, body mass index (BMI), and CD4 count.

### Analyses

Because creatinine had a right skewed distribution, it was log transformed to shift the distribution toward normality. Continuous variables were summarized as means and reported with standard deviations (SDs), skewness, kurtosis, medians, quartiles and ranges and categorical variables were reported as percentages. Time on TDF at the post-TDF creatinine measure was calculated as the difference between the date of the post-TDF initiation creatinine measurement and the date of TDF initiation. We evaluated eGFR, log-transformed creatinine, and pre-to post-TDF initiation changes in these values as continuous variables. T-tests and nonparametric tests compared continuous baseline characteristics while chi-square tests compared categorical characteristics among groups of patients. All adults initiating TDF in the study period defined, based on pre and post creatinine measurements they had have been recorded. Paired t-test and sign tests compared changes from baseline to follow up for continuous variables and McNemar’s discordant pairs tests did so for binary variables.

## Results

Among 476 patients included in the analysis, mean age at initiation of TDF was 35.9 years (SD 9.6); mean BMI was 22.0 kg/m^2^ (SD 4.1) and mean CD4 count was 544 (SD, 310) cells/μL (**Table 1**). The post-TDF creatinine test was measured at a mean of 95 days after TDF initiation. Mean pre- and post-TDF creatinine measurements were 0.94 (SD 0.33 mg/dl) and 0.89 (SD 0.20 mg/dl) respectively, corresponding to a pre-to post-TDF decline in creatinine of 0.055 mg/dL (P= 0.006). Mean pre-TDF eGFR was 92.4 (SD 24.0) and mean post-TDF was 96.0 (SD, 21.0) mL/min/1.73m^2^, corresponding to an increase of 3.6 (SD 26.6) mL/min/1.73m^2^ (p=0.001).

**Table 1.** Demographic characteristics of the study sample and changes in creatinine and eGFR^a^ from before to after TDF initiation^b^.

| Variable | Mean (SD) or Percentage<br>N = 476 |
| --- | --- |
| **DEMOGRAPHIC CHARACTERISTICS** |  |
| Percent women | 55.5% |
| Age at TDF Initiation, years | 35.9 (9.6) |
| BMI (kg/m<sup>2</sup>) | 22.0 (4.1) |
| CD4 (cells/μL) | 544 (310) |
| **PRE- POST- TDF CREATININE / EGFR AND CHANGES** |  |
| Pre-Creatinine (mg/dl) | 0.94 (0.33) |
| Post-Creatinine (mg/dl) | 0.89 (0.20) |
| Pre-post TDF Change in creatinine<sup>c</sup> | -0.055 (0.357) |
| Pre-eGFR (mL/min/1.73m<sup>2</sup>) | 92.4 (24.0) |
| Post-eGFR (mL/min/1.73m<sup>2</sup>) | 96.0 (21.0) |
| Pre-post TDF Change in eGFR<sup>d</sup> | 3.60 (26.62) |
| Duration of TDF use at testing, days | 95 (73) |

a. Estimated Glomerular Filtration Rate calculated by CKD-EPI equation without race adjustment; b. Measurement after TDF done less than 12 months after initiation. c. P-value for mean and median change to be 0 is 0.006 by paired t-test and 0.002 by signed rank test. d. P-value for mean and median change to be 0 is 0.001 by paired t-test and 0.005 by signed rank test.

When analyzing pre-to post-TDF creatinine and eGFR changes stratified by sex, age, CD4 and BMI, there were no qualitative or statistical differences among the subgroups (data not shown). Only two patients had stage 4 or 5 renal disease (defined as eGFR <30 mL/min/1.73m^2^) at the pre-TDF measure. No patients including these two had stage 4 or 5 renal disease at the post-TDF measure.

## Discussion

In this cohort of 476 patients receiving routine HIV care at 10 Rwandan health centers after national implementation of Treat All, we found no evidence that TDF was associated with impaired renal function during the first six months of administration, and conversely observed a small but statistically significant increase in eGFR after TDF initiation. This adds to the small literature on the association between TDF and renal function in SSA, suggesting that TDF can continue to be safely used as ART is expanded to all PLWH in this region.

We found no evidence of TDF-associated renal dysfunction among patients included in this analysis These results are somewhat different than those from the few published studies from other SSA countries, which reported a low but substantial incidence of TDF-associated renal dysfunction. In a Ugandan study, severe kidney dysfunction (eGFR<30 mL/min by Cockcroft-Gault) occurred in 2.7% of patients on all ARV regimens(1), with TDF the drug most often associated with Fanconi syndrome, a rare disorder of kidney tubular function(1). A Zambian study reported a low prevalence of TDF-induced renal insufficiency (i.e. eGFR < 50 mL/min) at six months (2.6%) and 12 months (3.1%) after TDF initiation (8). Our study, as other small literature in SSA, is supportive of TDF use. Nevertheless, our results might be different for some specific reasons, including a clinically healthier study population as evidence by a mean CD4 value of 544±310 cells/μL, (P-Value: 0.008), and shorter follow up time.

Our study had some limitations. First, pre- and post-TDF creatinine measures were available for only 31% of all patients who initiated TDF. However, the higher baseline creatinine among those with both pre- and post-TDF measurements compared to those with pre-TDF measurements only suggests that patients with worse renal function may be selectively targeted for creatinine measurements (See supplemental appendix). If so, then selective exclusion of patients with better renal function may mean that our results underestimate the post-TDF increase in eGFR among all PLWH initiating TDF. Another limitation is the follow-up of only six months, which may not have provided sufficient time to detect decreases in eGFR associated with TDF use.

TDF is currently recommended as a first line agent in combination with other ARVs for treatment of HIV-infection. This is one of the first studies to look at changes in renal function after TDF initiation in SSA under Treat All. We detected a statistically significant, but clinically small, improvement in mean renal function after 6 months of TDF use among 476 ART-naïve patients initiating TDF-based first line ART after July 1, 2016. While some studies outside of Africa have found renal toxicity in PLWH on TDF(1)(3)(4), our findings are in agreement with most of the studies from Africa and outside of Africa that have reported TDF to be associated with good clinical outcomes and found to have few side effects in healthy patients(9)(13)(14)(15)(16)

As Treat All continues to be implemented across SSA, our findings support the current WHO guidelines that advise the regular use of TDF regimens for all PLWH without known renal dysfunction, with serum creatinine monitoring for potential deterioration in renal function.

## Data Availability

Data used in this study are available on the data set of the Central Africa International Epidemiologic Databases to Evaluate AIDS (IeDEA)

https://www.iedea.org

## Disclaimer

The findings and conclusions in this report are those of the author(s) and have no conflict of interest to declare.

## Notes

### Competing Interest Statement

The authors have declared no competing interest.

### Clinical Trial

This is a research article, not a clinical trial

### Funding Statement

There is no sternal funding to report

### Author Declarations

This study was approved by the Rwanda National Ethics Committee and the institutional review board of the Albert Einstein College of Medicine

## References

1. Kalyesubula R, Perazella MA. Nephrotoxicity of HAART. AIDS Res Treat. 2011;2011.

2. Agbaji OO, Abah IO, Ebonyi AO, Gimba ZM, Abene EE, Gomerep SS, et al. Long Term Exposure to Tenofovir Disoproxil Fumarate-Containing Antiretroviral Therapy Is Associated with Renal Impairment in an African Cohort of HIV-Infected Adults. J Int Assoc Provid AIDS Care. 2019;18:1–9.

3. Fritzsche C, Rudolph J, Huenten-Kirsch B, Hemmer CJ, Tekoh R, Kuwoh PB, et al. Effect of tenofovor diproxil fumarate on renal function and urinalysis abnormalities in HIV-infected Cameroonian adults. Am J Trop Med Hyg. 2017;97(5):1445–50.

4. Quesada PR, Esteban LL, Garcia JR, Sanchez RV, Garcia TM, Alonso-Vega GG, et al. Incidence and risk factors for tenofovir-associated renal toxicity in HIV-infected patients. Int J Clin Pharm. 2015;37(5):865–72.

5. Ryom L, Mocroft A, Kirk O, Worm SW, Kamara DA, Reiss P, et al. Association between antiretroviral exposure and renal impairment among HIV-positive persons with normal baseline renal function: The D:A:D Study a. J Infect Dis. 2013;207(9):1359–69.

6. Jose S, Hamzah L, Campbell LJ, Hill T, Fisher M, Leen C, et al. Incomplete reversibility of estimated glomerular filtration rate decline following tenofovir disoproxil fumarate exposure. J Infect Dis. 2014;210(3):363–73.

7. De Waal R, Cohen K, Fox MP, Stinson K, Maartens G, Boulle A, et al. Changes in estimated glomerular filtration rate over time in South African HIV-1-infected patients receiving tenofovir: a retrospective cohort study. J Int AIDS Soc [Internet]. 2017;20(1):21317. Available from: https://doi.org/10.7448/IAS.20.01/21317

8. Wandeler G, Keiser O, Mulenga L, Christopher J. Collaborative analysis of cohort studies. 2013;61(1):41–8.

9. Cooper RD, Wiebe N, Smith N, Keiser P, Naicker S, Tonelli M. Systematic Review and Meta-analysis: Renal Safety of Tenofovir Disoproxil Fumarate in HIV-Infected Patients. Clin Infect Dis. 2010;51(5):496–505.

10. Nash D, Yotebieng M, Sohn AH. Treating all people living with HIV in sub-Saharan Africa: a new era calling for new approaches. J virus Erad [Internet]. 2018;4(Suppl 2):1–4. Available from: http://www.ncbi.nlm.nih.gov/pubmed/30515307%0Ahttp://www.pubmedcentral.nih.gov/articlerender.fcgi?artid=PMC6248848

11. NISR. Rwanda Demographic and Health Survey 2014-15 - Final Report. Rwanda. 2015. 640 p.

12. Rwanda Ministry of Health. National Guidelines for Prevention and Management of HIV and STIs. Edition 201 6. 2016;38.

13. Chi BH, Mwango A, Giganti M, Mulenga LB, Tambatamba-Chapula B, Reid SE, et al. Early clinical and programmatic outcomes with tenofovir-based antiretroviral therapy in Zambia. J Acquir Immune Defic Syndr. 2010;54(1):63–70.

14. Badii VS, Buabeng KO, Agyarko Poku T, Forkuo AD, Boamah BB, Arhin SM, et al. Tenofovir-Based Highly Active Antiretroviral Therapy Is Associated with Superior CD4 T Cells Repopulation Compared to Zidovudine-Based HAART in HIV 1 Infected Adults. Int J Chronic Dis. 2018;2018:1–8.

15. Siberry GK, Williams PL, Mendez H, Iii GRS, Jacobson DL, Hazra R, et al. NIH Public Access. 2013;26(9):1151–9.

16. Onoya D, Hirasen K, van den Berg L, Miot J, Long LC, Fox MP. Adverse Drug Reactions Among Patients Initiating Second-Line Antiretroviral Therapy in South Africa. Drug Saf [Internet]. 2018;41(12):1343–53. Available from: https://doi.org/10.1007/s40264-018-0698-3

